# Distinct temporal characteristics of circulating alveolar epithelial and endothelial injury markers in ARDS with COVID-19: a preliminary retrospective report

**DOI:** 10.1101/2021.01.10.21249528

**Authors:** Kentaro Tojo, Natsuhiro Yamamoto, Takahiro Mihara, Miyou Abe, Takahisa Goto

**Affiliations:** Department of Anesthesiology and Critical Care Medicine, Yokohama City University School of Medicine, Yokohama, Kanagawa, Japan; Department of Health Data Science, Yokohama City University Graduate School of Data Science, Yokohama, Kanagawa, Japan

## Abstract

The time course and specific contributions of alveolar epithelial and endothelial injury to the pathogenesis of acute respiratory distress syndrome (ARDS) with coronavirus disease (COVID-19) remain unclear. Here, we evaluated the characteristics of circulating markers of alveolar epithelial and endothelial injury in serum samples from eleven ARDS patients and ten non-ARDS patients, all with COVID-19. Our results indicates that the alveolar epithelial injury at the very early disease stage and the endothelial injury which continues to exacerbate during the later disease stage seem to be the hallmarks of ARDS with COVID-19.

## Background

In the most severe cases, Coronavirus disease (COVID-19) leads to acute respiratory distress syndrome (ARDS) that is characterized by alveolar epithelial and endothelial injuries [1,2]. Evaluating the temporal changes of established alveolar tissue injury markers[3] can provide insights regarding the time course and specific contribution of alveolar epithelial and of endothelial injury to the pathogenesis of COVID-19 ARDS. Recently, Spadaro et al. have reported that COVID-19 ARDS is characterized by the increases of circulating endothelial injury markers[4]. However, the detailed temporal characteristics of changes in these markers are still not clear. In this preliminary study, we investigated the levels of a circulating alveolar epithelial injury marker; soluble receptor for advanced glycation end-products (sRAGE), an endothelial injury marker; angiopoietin-2 (ANG-2), and an alveolar barrier permeability indicator; surfactant protein D (SP-D) in serum from COVID-19 patients with or without ARDS.

## Methods

The patients diagnosed as COVID-19 by real-time polymerase chain reaction and admitted to Yokohama City University Hospital from January to August 2020 were included in this retrospective observational study (ethics reference number: B200700100). Concentrations of sRAGE, ANG-2, and SP-D in the residual serum samples were measured using enzyme-linked immunosorbent assay kits (human RAGE: DY1145, human ANG-2: DY623, human SP-D: DY1920, R&D systems, MN, USA). We compared the concentrations of these markers on the first or second hospital day between ARDS and non-ARDS patients. Moreover, we analyzed temporal changes of these markers during the first eight hospital days in ARDS patients.

The data between ARDS and non-ARDS patients were compared with the Mann-Whitney U test. Temporal changes of the markers were analyzed with Friedman test and post-hoc Dunn’s test. The peak day of the each of the markers were analyzed with Kruskal-Wallis test and post-hoc Dunn’s test. All statistical analyses were performed using Prism 9.0 software (Graphpad Software, CA, USA). Statistical significance level was set at P < 0.05.

## Results

Eleven ARDS and ten non-ARDS patients, all with COVID-19, were included in the study. Patient characteristics are shown in Table 1. The diagnosis of ARDS was made on the first or second hospital day. The initial serum levels of sRAGE and SP-D were significantly higher in the ARDS group than in the non-ARDS group, however the ANG-2 level was not (Table.1).

**Table.1.**
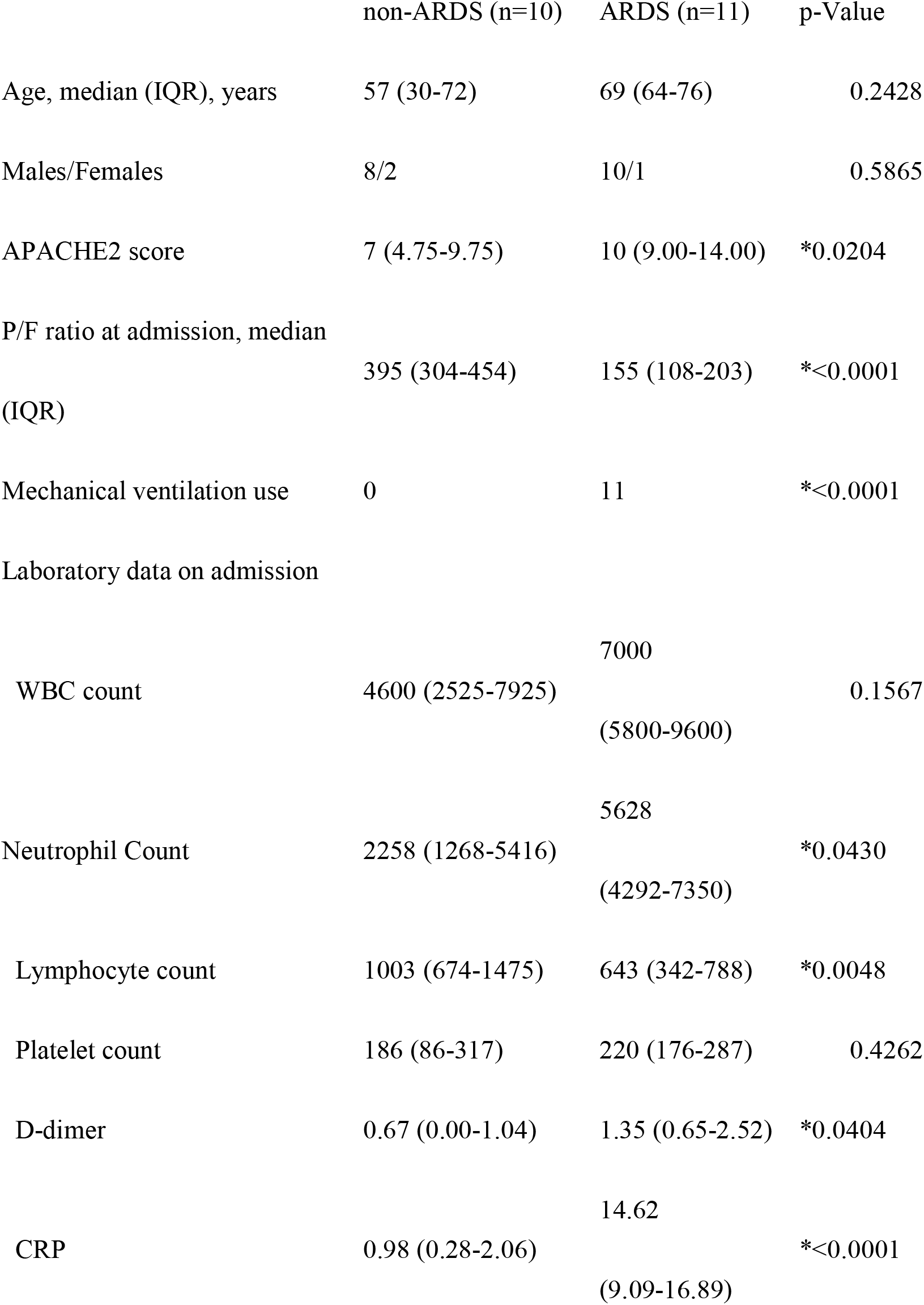
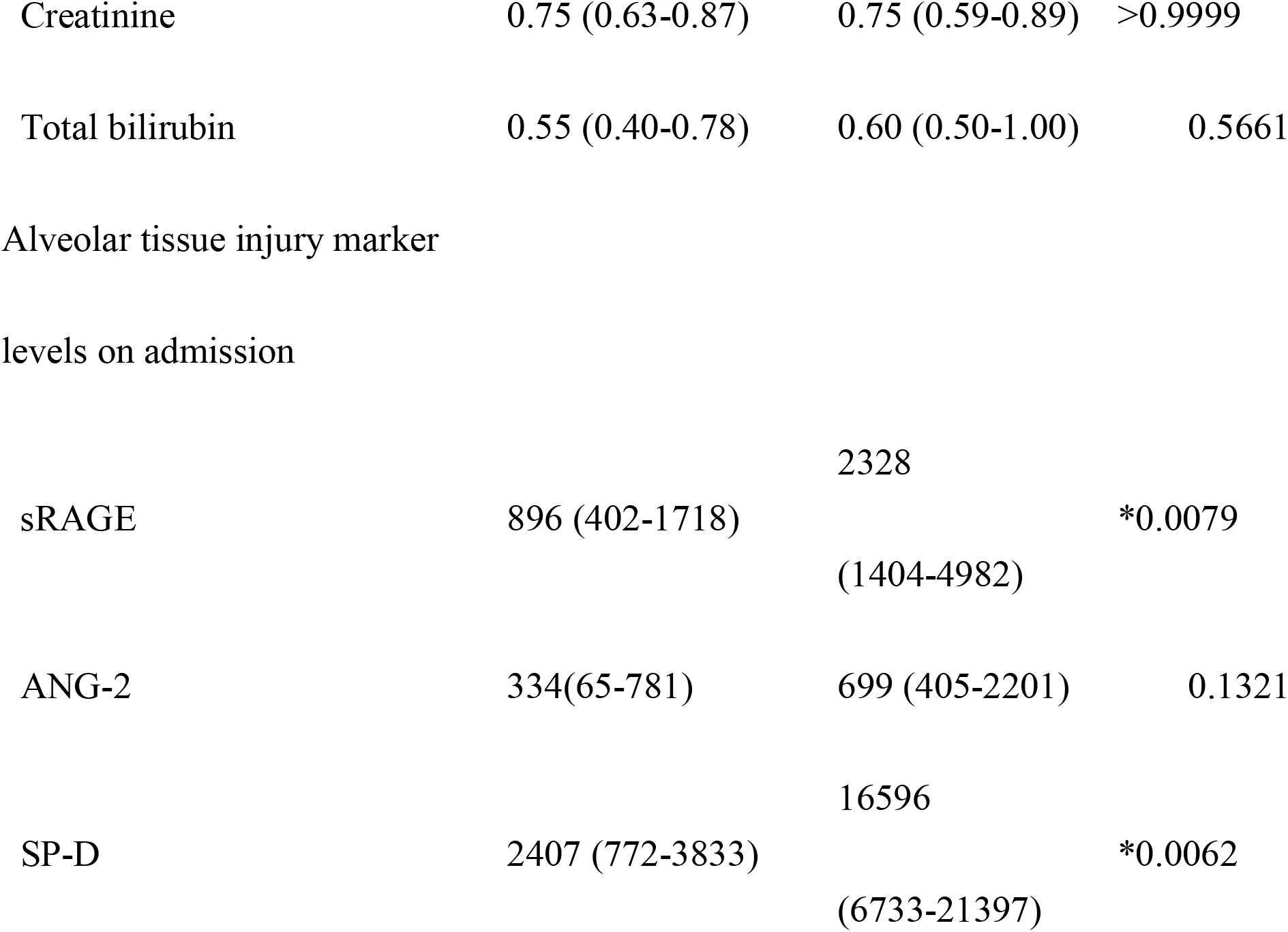

Analysis of the temporal changes of these markers in ten ARDS patients excluding one patient with missing data revealed that serum sRAGE level peaked just after admission, and it gradually decreased with hospital days (Fig.1). In contrast, serum ANG-2 and SP-D did not significantly decrease during first eight hospital days and the peak timings of these markers were during a later disease stage (Fig.1).

**Figure.1.**
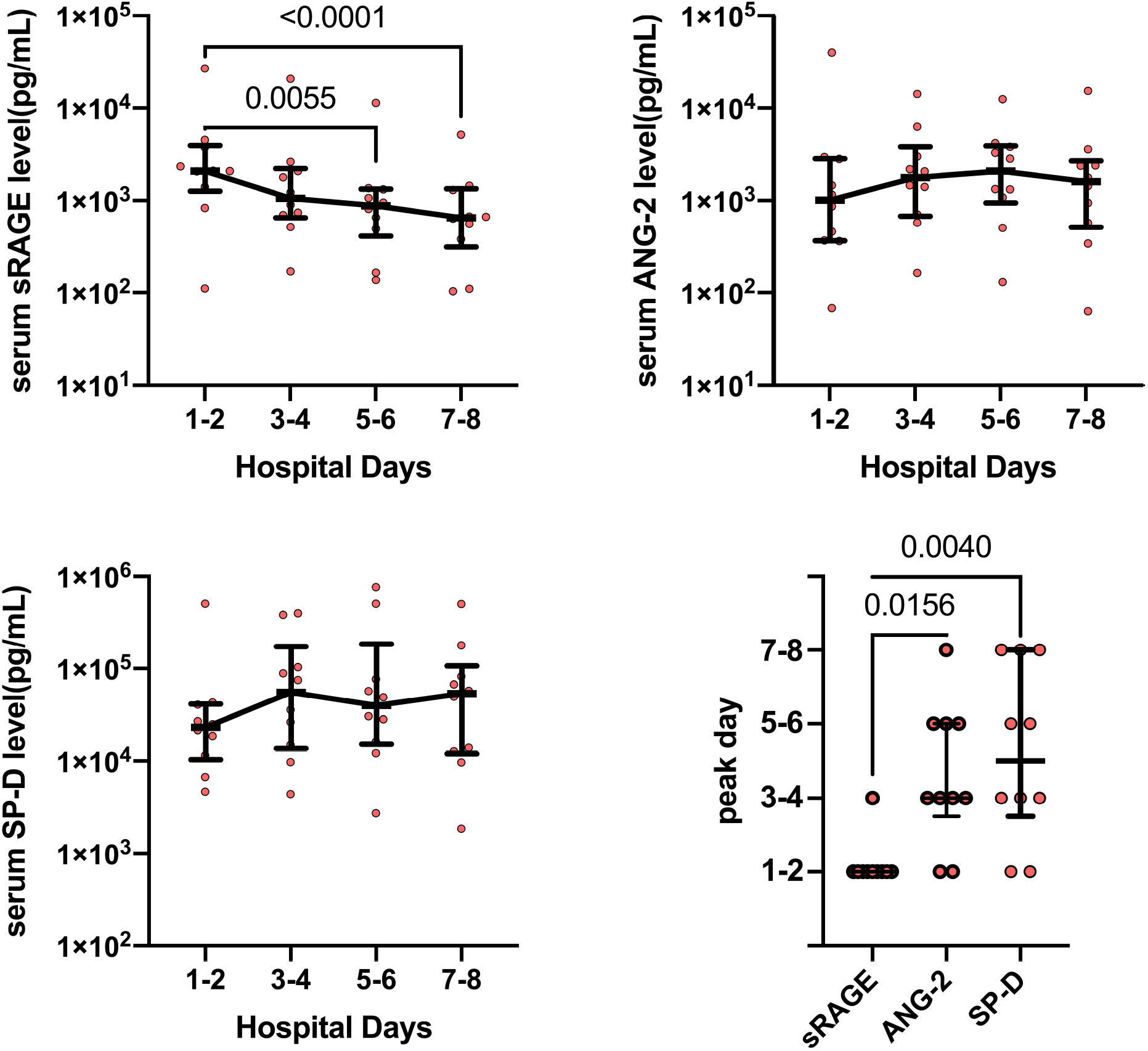
Temporal changes of (A) sRAGE, (B) ANG-2, and (C) SP-D levels in ARDS patients with COVID-19 for 8 days starting from admission. In cases where values for every 2-days were available, then an average of the values was used; where only a single day value was available, then that was used. (D) The peak day of each of the alveolar tissue injury markers. Data were presented as median ± IQR.

## Discussion

The present study indicates that alveolar epithelial injury occurring at the very early disease stage, which is indicated by the increase in sRAGE level, is a hallmark of COVID-19 ARDS pathogenesis. On the other hand, levels of ANG-2 peaked at later time points, suggesting that the endothelial injury continue to exacerbate for several days after admission. Moreover, the trajectory of ANG-2 level, rather than of sRAGE level, was parallel with the trajectory of SP-D, an indicator of alveolar barrier permeability. These data suggest that the endothelial injury contribute to the exacerbation of the alveolar barrier disruption during the later disease stage.

The difference in the peak timing of epithelial and endothelial injury markers suggests the distinct mechanisms for injury to each type of cell. A hypothesis generated from our results is that the initial alveolar epithelial injury might be a trigger of the subsequent endothelial injury. To obtain insights into the pathogenesis of alveolar tissue injury during COVID-19, further studies are warranted.

## Data Availability

De-identified patient data collected for the study will be rendered available by writing to KT.

## List of abbreviations

ARDS: acute respiratory distress syndrome
COVID-19: coronavirus disease
sRAGE: soluble receptor for advanced glycation end-products
ANG-2: angiopoietin-2
SP-D: surfactant protein D
SARS-CoV-2: severe acute respiratory syndrome coronavirus-2

## Declarations

### Ethics approval and consent to participate

The study protocol was reviewed and approved by the institutional review board of Yokohama City University Hospital (approval number: B200700100). The need for informed consent was waived by the institutional review boards because of the retrospective observational design of the study.

### Consent for publication

Not applicable.

### Availability of data and materials

The datasets used and/or analysed during the current study are available from the corresponding author on reasonable request.

### Authors’ contributions

KT conducted the study, performed ELISA, analysed data, and wrote the manuscript. NY performed ELISA and reviewed the manuscript. TM supervised statistical data analysis and reviewed the manuscript. MA collected patients’ clinical data and reviewed the manuscript. TG supervised the study and reviewed the manuscript

## Acknowledgements

The authors wish to thank Department of Emergency Medicine (Prof. Ichiro Takeuchi), Department of Microbiology (Prof. Akihide Ryo), and Yokohama City University Center for Novel and Exploratory Clinical Trials for collecting and providing the blood samples.

